# Epigenome-wide association study using prediagnostic bloods identifies a new genomic region (near *TMEM204 and IFT140*) associated with pancreatic cancer risk

**DOI:** 10.1101/2020.02.07.20021121

**Authors:** Dominique S. Michaud, Mengyuan Ruan, Devin C. Koestler, Dong Pei, Carmen J. Marsit, Immaculata De Vivo, Karl T. Kelsey

## Abstract

**Background:** Epigenome-wide association studies (EWAS) using peripheral blood have identified specific sites of DNA methylation associated with risk of various cancers and may hold promise to identify novel biomarkers of risk; however, few studies have been performed for pancreatic cancer and none using a prospective study design.

**Methods:** Using a nested case-control study design, incident pancreatic cancer cases and matched controls were identified from participants who provided blood at baseline in three prospective cohort studies (Nurses’ Health Study, Health Professionals Follow-up Study and Physicians’ Health Study). DNA methylation levels were measured in DNA extracted from leukocytes using the Illumina MethylationEPIC array. Average follow-up period for this analysis was 13 years.

**Results:** A region in chromosome 16 near genes*TMEM204 and IFT140* was identified as being differentially methylated in cases and controls. For some CpGs in the region, the associations were stronger with shorter time to diagnosis (e.g., OR= 5.95, 95% CI = 1.52-23.12, for top vs bottom quartile, for <5 years between blood draw and cancer diagnosis) but associations remained significantly higher even when cases were diagnosed over 10 years after blood collection. Statistically significant differences in DNA methylation levels were also observed in the gastric secretion pathway using GSEA analysis.

**Conclusions:** Changes in DNA methylation in peripheral blood may mark alterations in metabolic or immune pathways (potentially including alterations in immune subtypes) that play a role in pancreatic cancer. Identifying new biological pathways in carcinogenesis of pancreatic cancer using EWAS approach could provide new opportunities for improving treatment and prevention.

## Introduction

Pancreatic cancer accounts for the third highest number of cancer-related deaths in the US, after lung and colorectal cancers, and is expected to surpass colorectal cancer in the next decade [1]. Pancreatic cancer is highly lethal, with 93% of pancreatic cancer patients succumbing to their disease within 5 years of diagnosis, largely due to most cases being diagnosed at late stages. Screening for pancreatic cancer is not currently recommended for asymptomatic adults as existing screening methods have not been shown to reduce mortality [2]. Improving sensitivity of tests is important, but for screening to be effective patients need to be identified at early stages of the disease. Moreover, because pancreatic incidence rates are low, identifying high risk groups for screening is necessary to increase the positive predictive value of the screening tests.

Some studies aimed at identifying early detection biomarkers have focused on measuring DNA methylation of promoter regions in known cancer genes using peripheral cell-free (cf)DNA in bloods of patients [3, 4]. These methods are promising but it is unclear whether they would identify cancers at early stages of the disease, when treatment would be most effective. Alternatively, high-dimensional arrays now provide the opportunity to agnostically interrogate 100s of thousands of biomarkers, including quantification of DNA methylation throughout the genome. Using these technologies, studies have begun to examine how DNA methylation levels in peripheral leukocytes vary by cancer status [5-7].

Several case-control studies on pancreatic cancer have compared DNA methylation in blood leukocytes of pancreatic cancer patients with healthy controls [8-10]. However, these studies were retrospective case-control studies and they cannot differentiate between recent changes to those that occurred months or years prior to diagnosis. Conducting prospective nested case-control studies (i.e., selected subjects from existing cohort studies), using blood samples that were obtained from healthy individuals many years prior to diagnosis, provide a unique opportunity to identify biomarkers that may be linked to disease progression. As DNA methylation levels can change over time to impact gene expression, identifying changes decades prior to disease could assist in uncovering pathways that contribute to the development of cancer, and novel genomic regions identified using these methods could potentially be targeted for prevention or drug intervention.

To our knowledge, this study presents results from the first epigenome-wide association study (EWAS) on pancreatic cancer risk using pre-diagnostic peripheral blood leukocytes. For this study, 393 pancreatic cancer cases and matched 431 controls were obtained from three large cohort studies where blood samples had been collected between 6 months and 26 years prior to cancer diagnosis.

## Methods

The primary EWAS was conducted using prediagnostic bloods of pancreatic cancer cases and matched controls selected from the Nurses’ Health Study (NHS), the Physicians’ Health Study (PHS), and the Health Professionals Follow-up Study (HPFS).

For this analysis, 403 incident cases were confirmed to have pancreatic cancer among the participants who provided blood samples prior to cancer diagnosis. A control subject was matched to each case on cohort (which also matches on sex), age (+/- 1 year), date of blood draw (month 3+/- and year), smoking (never, former, current) and race (White/other). Due to low DNA concentrations in some of the samples, and samples removed after data processing of the arrays (see Supplementary Methods), the initial 1:1 matching was not always conserved and resulted in some cases and controls with no matched pair. The final dataset consisted of 393 cases and 431 controls.

### DNA methylation measurements

DNA extracted from buffy coats were bisulfite-treated and DNA methylation was measured with the Illumina Infinium MethylationEPIC BeadChip array (Illumina, Inc, CA, USA). Details on DNA methylation measurements and data processing are provided in the Supplementary Methods.

### Statistical analyses

All statistical analyses were performed in R (version 3.5.1). The dataset of cases and controls from the cohort studies was randomly divided into a training set (N=577) and testing set (N=247). We initially conducted the EWAS analysis using a series of unconditional multivariable logistic regression models to examine the association between the CpG-specific DNA methylation and pancreatic cancer risk. Unconditional logistic regression models were used to maximize power by including cases and controls without matched pairs. All models for the

EWAS analysis were adjusted for age at blood draw, cohort, race (white/non-white), smoking status (never, former, current), date of blood draw (continuous), BMI, and cell composition [11, 12], given the potential for confounding by cell composition [13]. Models were fit to the training and testing sets separately, with the intent to compare significant results. All p-values were adjusted for multiple comparisons using a Bonferroni correction in the EWAS. Statistical method section for the risk prediction score analysis is provided in the Supplemental Methods.

New statistical methods, such as DMRcate [14], have been developed to test for regions that are differentially methylated (DMRs) by phenotype using DNA methylation arrays; we used the DMRcate Bioconductor R package to identify DMRs associated with pancreatic cancer risk. DMRcate was applied independently to the training and testing sets using the following settings: region length 2000bp; min of 2 significant CpGs; FDR pvalue<0.05 (the same covariates were adjusted for as in the EWAS). Regions identified that were statistically significant after correction for multiple comparisons were compared between the training and testing sets. CpGs identified in regions that were identified as statistically significant in both datasets were further evaluated by using the combined dataset to test associations with pancreatic cancer risk using CpG quartiles (based on methylation levels in the controls). Separately, we examined associations by cohort (using cohort-specific cutpoints for the quartiles) and by time from blood-draw to diagnosis.

Because different cell types might exhibit differing associations between CpG-specific DNA methylation and pancreatic case control status, an additional analysis was conducted to identify CpGs that might be unique to different cell types in blood. To carry this out, we applied Tensor Composition Analysis (TCA)[15], a computational approach for estimating cell-specific methylation signatures based on bulk DNA methylation data (e.g., whole-blood). Cell-specific methylation profiles were estimated for CD4T, CD8T, NK, B cell, monocytes, and neutrophils across all study samples, and used to test for cell-specific effects of methylation on pancreatic cancer risk. Unconditional multivariable logistic regression models adjusted for age at blood draw, cohort, race, smoking status (never, former, current), date of blood draw (continuous), and BMI were used to examine the cell-specific and CpG-specific association between DNA methylation and pancreatic cancer risk. The p-values from the GSEA were FDR adjusted.

To identify biological pathways and gene ontologies associated with pancreatic case control status, we used the methylGSA Bioconductor package [16] to perform gene set analyses. Analyses were performed separately on the training and testing sets and the top gene sets (rank-ordered based on p-value) were compared between the training and testing sets to look for consistent pathways/ontologies (models adjusted for age at blood draw, cohort, smoking status (never, former, current), date of blood draw (continuous), and BMI).

## Results

The population characteristics for the cases and controls overall, and by the training and testing sets, are presented in Table 1. Participants in the nested case-control study were diagnosed with pancreatic cancer an average of 13 years (range 6 months to 26 years) after providing a blood sample. About 45% of cases/controls were women (selected from the NHS cohort), and the remaining cases/controls were men from the PHS (18/20%) and HPFS (37/35%) cohorts. Baseline characteristics were evenly distributed between the training and testing datasets (Table 1).

**Table 1.** Baseline characteristics * for subjects in three prospective cohort studies (NHS, HPFS, PHS) included in the nested case-control analysis.

| Mean (SD) or N (%) | **Total (N = 824)** |  | **Training (N = 577)** |  | **Testing (N = 247)** |  |
| --- | --- | --- | --- | --- | --- | --- |
|  | Cases | Controls | Cases | Controls | Cases | Controls |
| **N** | 393 | 431 | 276 | 301 | 117 | 130 |
| **Cohort** |  |  |  |  |  |  |
| NHS | 176 (44.8%) | 194 (45.0%) | 126 (45.7%) | 137 (45.5%) | 50 (42.7%) | 57 (43.8%) |
| HPFS | 146 (37.2%) | 151 (35.0%) | 103 (37.3%) | 105 (34.9%) | 43 (36.8%) | 46 (35.4%) |
| PHS | 71 (18.1%) | 86 (20.0%) | 47 (17.0%) | 59 (19.6%) | 24 (20.5%) | 27 (20.8%) |
| **Age at blood draw** | 60.6 (7.9) | 60.2 (7.7) | 61.0 (8.1) | 60.4 (7.8) | 59.8 (7.5) | 59.8 (7.5) |
| **Time before diagnosis (year)<sup>†</sup>** | 13.0 (6.2) |  | 13.2 (6.2) |  | 12.4 (6.2) |  |
| **Female** | 176 (44.8%) | 194 (45.0%) | 126 (45.7%) | 137 (45.5%) | 50 (42.7%) | 57 (43.8%) |
| **White<sup>‡</sup>** | 346 (94.0%) | 406 (94.9%) | 244 (93.8%) | 284 (95.3%) | 102 (94.4%) | 122 (93.8%) |
| **Smoking<sup>§</sup>** |  |  |  |  |  |  |
| Never | 159 (40.9%) | 173 (40.3%) | 111 (40.5%) | 118 (39.5%) | 48 (41.4%) | 55 (42.6%) |
| Former | 174 (44.7%) | 190 (44.3%) | 120 (43.8%) | 134 (44.8%) | 54 (46.6%) | 56 (43.4%) |
| Current | 57 (14.6%) | 65 (15.2%) | 43 (15.7%) | 47 (15.7%) | 14 (12.1%) | 18 (14.0%) |
| **BMI (kg/m<sup>2</sup>)** | 26.0 (4.3) | 25.6 (3.9) | 26.1 (4.3) | 25.5 (3.9) | 25.9 (4.1) | 25.9 (4.0) |
| Underweight and normal | 183 (47.3%) | 215 (50.8%) | 132 (48.7%) | 155 (52.7%) | 51 (44.0%) | 60 (46.5%) |
| Overweight | 144 (37.2%) | 151 (35.7%) | 91 (33.6%) | 100 (34.0%) | 53 (45.7%) | 51 (39.5%) |
| Obese | 60 (15.5%) | 57 (13.5%) | 48 (17.7%) | 39 (13.3%) | 12 (10.3%) | 18 (14.0%) |
| **Diabetes<sup>¶</sup>** | 19 (4.8%) | 11 (2.6%) | 14 (5.1%) | 10 (3.3%) | 5 (4.3%) | 1 (0.8%) |

SD: standard deviation, BMI: body mass index, NHS: Nurses' Health Study, HPFS: Health Professionals Follow-up Study, PHS: Physicians' Health Study.
\* Covariates based on questionnaires closest to time of blood draw.
<sup>†</sup> 11 missing values; <sup>‡</sup> 28 missing values; <sup>§</sup> 6 missing values; <sup>||</sup> 14 missing values; <sup>¶</sup> 1 missing values.

The single CpG EWAS analyses performed in the training and testing sets (conducted separately) identified no statistically significant CpGs after a Bonferroni correction. We reduced our number of comparisons by including only CpGs with ICC >0.5 (over a 1-year period) from our pilot study (n=199,719 CpGs)[17], but still no CpGs were statistically significant using this restrictive number of tests.

Using the training dataset, we identified 99 CpGs using our cutpoints for mean difference and statistical significance using a volcano plot analysis (Supplemental Figure 1). The LASSO regression model reduced that number of CpGs to 92. Before developing a polygenic risk score, we evaluated whether the mean difference in methylation in these CpGs was consistent in the training and testing datasets. Unfortunately, the mean difference was in different directions for 42 out of 92 CpGs, leading us to conclude that the polygenic risk score would not be able to discriminate between cases and controls in the testing set. Using only cases who had given blood within 5 years of diagnosis did not improve our ability to develop a risk prediction score.

### DMR analysis

To explore alternative approaches, we applied a method (DMRcate) designed to identify differentially methylated regions (DMRs) by case/control status [14]. Statistically significant regions were identified in each of the training and testing sets, one of which was statistically significant (and in the same direction) in both datasets; this region, located on chromosome 16 (1583391-1584516; overlapping promoters for *IFT140* and *TMEM204*), consisted of 13 CpGs that differed coordinately between the cases and controls in the two datasets (all were p<0.05 FDR adjusted) and an 3 additional CpGs in the testing dataset (Supplemental Tables 1&2). When we examined how these CpGs were associated by time between blood draw and cancer diagnosis there was a striking pattern of higher risk associated with time more proximal to diagnosis evident in many of the CpGs; results are presented in Table 2 and Figure 1. The strongest association was noted for cg09757087 (Table 3); a greater than 2-fold increase in risk was identified for individuals in the highest quartile of methylation level, compared to the lowest quartile, and a close to 6-fold increase was observed in subjects who provided blood <5 years prior to cancer diagnosis. For cg09757087, trends were significant across quartiles in all strata of time to diagnosis and in all but the PHS cohort study (Table 3). Of note, ICCs for the CpGs in this region calculated at two time points 1 year apart in a previous pilot study[17] were between 0.88 and 0.98.

**Table 2.** Top 12 CpGs in DMR region on chromosome 16 identified in training and testing datasets and combined for overall analysis.

| CpG | Annotation |  | Methylation Beta Value |  |  |  | Odds Ratio (Q4 vs. Q1)<sup>†</sup> |  |  |  |
| --- | --- | --- | --- | --- | --- | --- | --- | --- | --- | --- |
|  | Position | Relation to Island | Cases | Controls | SD | Diff / SD | Overall | Time to diagnosis |  |  |
|  |  |  |  |  |  |  |  | ≤ 5 years | 5-10 years | > 10 years |
| cg09757087* | 1585644 | S_Shore | 0.718 | 0.698 | 0.089 | 0.220 | 2.31(1.46, 3.66) | 5.92(1.52, 23.12) | 2.78(1.25, 6.19) | 2.21(1.30, 3.76) |
| cg11375102 | 1583810 | Island | 0.242 | 0.230 | 0.063 | 0.187 | 2.12(1.37, 3.26) | 3.90(1.34, 11.36) | 2.87(1.28, 6.42) | 1.76(1.07, 2.91) |
| cg27594616 | 1583620 | N_Shore | 0.218 | 0.205 | 0.081 | 0.160 | 1.89(1.25, 2.88) | 1.71(0.69, 4.28) | 2.11(0.99, 4.49) | 1.90(1.17, 3.09) |
| cg06565913 | 1584452 | Island | 0.368 | 0.352 | 0.086 | 0.184 | 1.86(1.22, 2.84) | 4.64(1.47, 14.64) | 3.19(1.45, 7.04) | 1.37(0.84, 2.24) |
| cg00977403* | 1585720 | N_Shore | 0.367 | 0.348 | 0.090 | 0.207 | 1.89(1.21, 2.94) | 2.45(0.96, 6.24) | 2.12(0.93, 4.80) | 1.82(1.08, 3.07) |
| cg16336651 | 1583391 | N_Shore | 0.542 | 0.531 | 0.056 | 0.197 | 1.75(1.16, 2.64) | 3.60(1.24, 10.50) | 2.76(1.31, 5.83) | 1.38(0.86, 2.21) |
| cg08296037 | 1584118 | Island | 0.393 | 0.372 | 0.095 | 0.215 | 1.75(1.15, 2.66) | 2.66(1.00, 7.07) | 2.43(1.10, 5.37) | 1.55(0.96, 2.51) |
| cg06602086 | 1583883 | Island | 0.297 | 0.278 | 0.111 | 0.173 | 1.74(1.14, 2.67) | 3.39(1.15, 9.99) | 2.53(1.15, 5.56) | 1.44(0.88, 2.34) |
| cg07341220 | 1583899 | Island | 0.204 | 0.192 | 0.075 | 0.163 | 1.72(1.13, 2.62) | 3.33(1.10, 10.06) | 2.47(1.13, 5.38) | 1.38(0.86, 2.21) |
| cg10465839 | 1584050 | Island | 0.259 | 0.246 | 0.079 | 0.165 | 1.70(1.12, 2.60) | 2.06(0.77, 5.49) | 2.58(1.18, 5.62) | 1.45(0.89, 2.36) |
| cg02193187 | 1583630 | N_Shore | 0.176 | 0.164 | 0.069 | 0.167 | 1.65(1.09, 2.51) | 1.85(0.75, 4.59) | 2.28(1.02, 5.06) | 1.53(0.94, 2.47) |
| cg07639376 | 1584516 | Island | 0.416 | 0.396 | 0.113 | 0.173 | 1.51(1.00, 2.28) | 1.85(0.75, 4.59) | 2.16(1.02, 4.57) | 1.33(0.83, 2.15) |

\*Only identified in testing dataset.
<sup>†</sup>Model adjusted for age at blood draw, race, date of blood draw, cohort study, smoking status, BMI and cell proportions.

**Figure 1.**
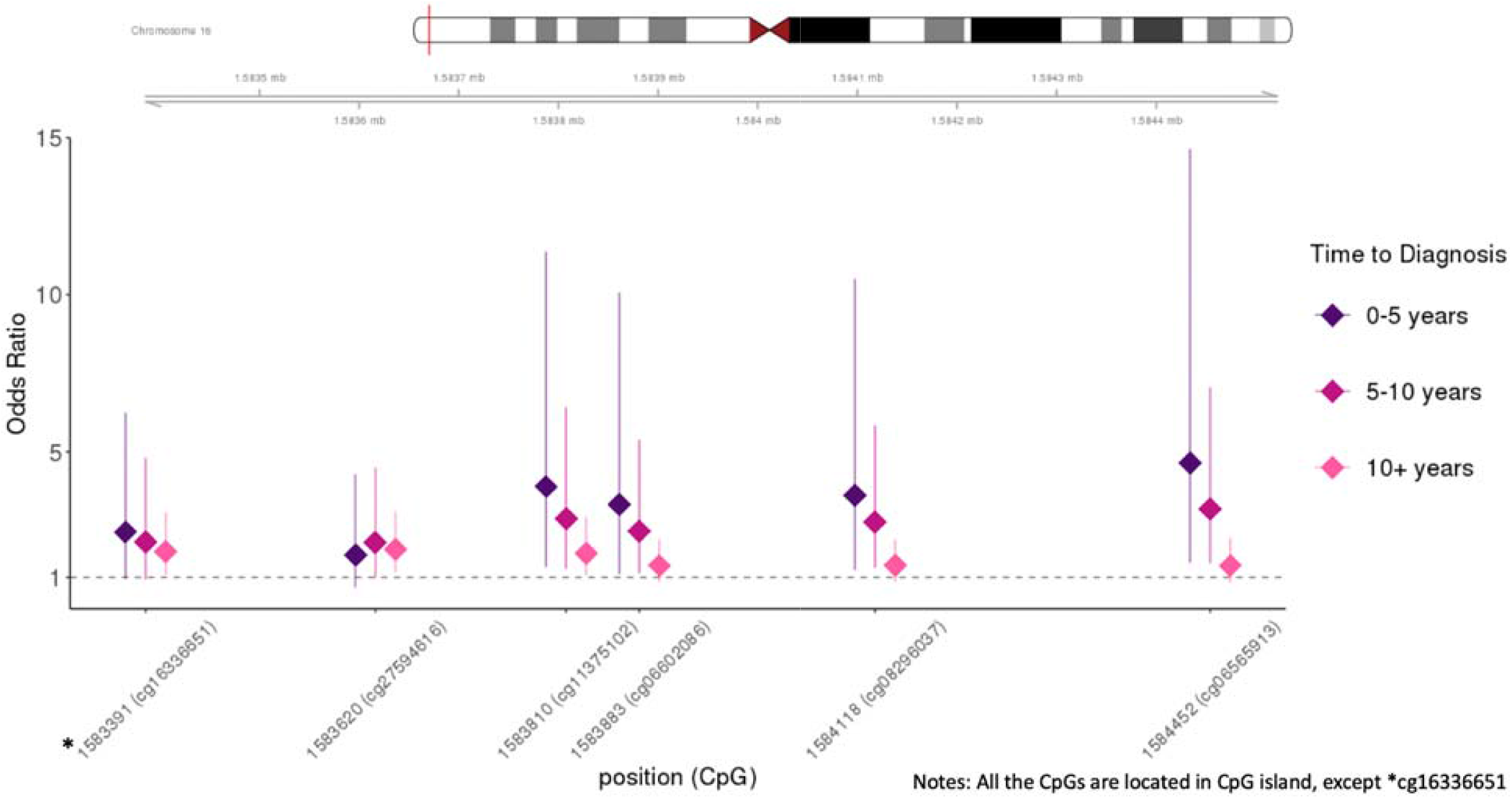
Odd ratios by time to diagnosis for top 6 CpGs in DMR region on chromosome 16 identified in both training and testing datasets. This figure only includes the CpGs that overlapped in training and testing dataset.

**Table 3.** Associations between cg09757087 and risk of pancreatic cancer across each quartile, overall, by time to diagnosis, and by cohort study

|  | **Model 1<sup>\*</sup>** |  | **Model 2<sup>†</sup>** |  |
| --- | --- | --- | --- | --- |
|  | Case / Control | OR (95% CI) | Case / Control | OR (95% CI) |
| **Overall (N = 824)<sup>‡</sup>** |  |  |  |  |
| **Q1** | 63 / 108 | ref. | 63 / 107 | ref. |
| **Q2** | 96 / 107 | 1.60(1.05, 2.44) | 93 / 103 | 1.61(1.05, 2.46) |
| **Q3** | 104 / 108 | 1.81(1.17, 2.79) | 99 / 104 | 1.76(1.13, 2.74) |
| **Q4** | 130 / 108 | 2.34(1.49, 3.68) | 129 / 107 | 2.31(1.46, 3.66) |
|  |  | p for trend <0.001 |  | p for trend <0.001 |
| **≤5 years to diagnosis<sup>‡</sup>** |  |  |  |  |
| **Q1** | 3 / 108 | ref. | 3 / 107 | ref. |
| **Q2** | 10 / 107 | 3.35(0.88, 12.71) | 10 / 103 | 3.39(0.89, 12.94) |
| **Q3** | 20 / 108 | 7.47(2.07, 26.97) | 19 / 104 | 7.47(2.04, 27.30) |
| **Q4** | 17 / 108 | 6.01(1.55, 23.24) | 17 / 107 | 5.92(1.52, 23.12) |
|  |  | p for trend = 0.004 |  | p for trend = 0.006 |
| **5-10 years to diagnosis<sup>‡</sup>** |  |  |  |  |
| **Q1** | 15 / 108 | ref. | 15 / 107 | ref. |
| **Q2** | 20 / 107 | 1.58(0.74, 3.38) | 20 / 103 | 1.54(0.71, 3.31) |
| **Q3** | 14 / 108 | 1.07(0.47, 2.45) | 13 / 104 | 0.95(0.40, 2.22) |
| **Q4** | 34 / 108 | 2.96(1.35, 6.48) | 33 / 107 | 2.78(1.25, 6.19) |
|  |  | p for trend = 0.01 |  | p for trend = 0.03 |
| **>10 years to diagnosis<sup>‡</sup>** |  |  |  |  |
| **Q1** | 41 / 108 | ref. | 41 / 107 | ref. |
| **Q2** | 62 / 107 | 1.60(0.98, 2.60) | 59 / 103 | 1.60(0.97, 2.62) |
| **Q3** | 69 / 108 | 1.93(1.17, 3.18) | 66 / 104 | 1.91(1.15, 3.19) |
| **Q4** | 77 / 108 | 2.16(1.28, 3.65) | 77 / 107 | 2.21(1.30, 3.76) |
|  |  | p for trend = 0.004 |  | p for trend = 0.004 |
| **NHS (N = 370)<sup>§</sup>** |  |  |  |  |
| **Q1** | 27 / 49 | ref. | 27 / 49 | ref. |
| **Q2** | 47 / 48 | 1.90(1.01, 3.60) | 47 / 48 | 1.94(1.02, 3.68) |
| **Q3** | 35 / 48 | 1.41(0.72, 2.77) | 33 / 47 | 1.37(0.69, 2.72) |
| **Q4** | 67 / 49 | 2.44(1.22, 4.90) | 67 / 49 | 2.49(1.24, 5.00) |
|  |  | p for trend = 0.04 |  | p for trend = 0.04 |
| **HPFS (N = 297)<sup>§</sup>** |  |  |  |  |
| **Q1** | 21 / 38 | ref. | 20 / 37 | ref. |
| **Q2** | 39 / 37 | 2.02(0.99, 4.14) | 38 / 34 | 2.23(1.06, 4.66) |
| **Q3** | 44 / 38 | 2.41(1.16, 5.00) | 40 / 34 | 2.63(1.22, 5.65) |
| **Q4** | 42 / 38 | 2.43(1.13, 5.23) | 41 / 37 | 2.67(1.20, 5.95) |
|  |  | p for trend = 0.03 |  | p for trend = 0.02 |
| **PHS (N = 157)<sup>§</sup>** |  |  |  |  |
| **Q1** | 16 / 22 | ref. | 16 / 22 | ref. |
| **Q2** | 13 / 21 | 0.98(0.36, 2.70) | 13 / 21 | 0.90(0.32, 2.53) |
| **Q3** | 25 / 21 | 1.71(0.68, 4.33) | 25 / 21 | 1.51(0.59, 3.89) |
| **Q4** | 17 / 22 | 1.47(0.49, 4.40) | 17 / 22 | 1.36(0.44, 4.16) |
|  |  | p for trend = 0.28 |  | p for trend = 0.38 |

<sup>\*</sup>Model 1: Adjusted for age at blood drawn, race, date of blood drawn, cohort study, and cell proportions.
<sup>†</sup>Model 2: Additional adjusted for smoking status and BMI.
<sup>‡</sup>Cut-offs (overall): Q1: ≤0.624; Q2: 0.625-0.703; Q3: 0.704-0.764; Q4: ≥0.765
<sup>§</sup>Cut-offs (study specific): Q1: $\leq 0.624$ for NHS, $\leq 0.625$ for HPFS and $\leq 0.622$ for PHS; Q2: 0.625-0.703 for NHS, 0.626-0.703 for HPFS and 0.626-0.701 for PHS; Q3: 0.704-0.764 for NHS, 0.704-0.765 for HPFS and 0.705-0.764 for PHS; Q4: $\geq 0.765$ for NHS, $\geq 0.766$ for HPFS and $\geq 0.766$ for PHS.

Although DNA methylation levels among control subjects in our study were similar across BMI and smoking categories, we conducted stratified analyses to examine whether our results were modified by BMI or smoking status. While the tests for interaction were not statistically significant, associations were stronger in overweight and obese subjects, and among current smokers (Supplemental Table 3). Results were similar among Whites only and those who did no report having diabetes at baseline (Supplemental Table 4).

We conducted additional analyses to identify DMRs that might be unique to different cell types in blood (see Methods for details on analytical procedures)[15]. Using this method, we were able to test whether the CpGs in the region identified with DMRcate with all cell types combined were differentially methylated in each of the immune cell types. Our results suggest that the methylation differences between cases and controls were not statistically significant in most cell types; however, we did observe strong associations for most of the CpGs in CD4 T cells and for 2 of the CpGs in CD8 T cells (Table 4).

**Table 4.** Top 5 CpGs (based on p-values) in the DMR region when examining associations for specific immune cell types*

|  | **Odds Ratio <sup>†</sup>**<br>**(Q4 vs. Q1)** | **95% CI** | **P-value** |
| --- | --- | --- | --- |
| **CD8T** |  |  |  |
| cg11375102 | 3.00 | (1.15, 7.80) | 0.024 |
| cg16336651 | 3.17 | (1.09, 9.18) | 0.033 |
| cg00463982 | 2.06 | (0.97, 4.36) | 0.061 |
| cg06602086 | 1.74 | (0.92, 3.29) | 0.091 |
| cg02193187 | 2.14 | (0.84, 5.43) | 0.110 |
| **CD4T** |  |  |  |
| cg10465839 | 2.50 | (1.26, 4.97) | 0.009 |
| cg02193187 | 1.81 | (1.11, 2.97) | 0.018 |
| cg00463982 | 2.20 | (1.11, 4.34) | 0.023 |
| cg08296037 | 2.83 | (1.11, 7.18) | 0.029 |
| cg11375102 | 1.94 | (1.07, 3.52) | 0.030 |
| **NK** |  |  |  |
| cg06602086 | 1.65 | (1.02, 2.69) | 0.042 |
| cg07639376 | 1.52 | (0.95, 2.41) | 0.078 |
| cg00463982 | 1.56 | (0.95, 2.58) | 0.078 |
| cg02193187 | 1.74 | (0.88, 3.45) | 0.110 |
| cg16336651 | 2.33 | (0.83, 6.59) | 0.110 |
| **Neutrophil** |  |  |  |
| cg16336651 | 1.76 | (1.13, 2.73) | 0.012 |
| cg27594616 | 1.61 | (1.06, 2.43) | 0.025 |
| cg26390081 | 1.50 | (1.00, 2.26) | 0.051 |
| cg00463982 | 1.47 | (0.97, 2.24) | 0.068 |
| cg06602086 | 1.46 | (0.96, 2.21) | 0.077 |
| **Monocytes** |  |  |  |
| cg06565913 | 1.50 | (0.97, 2.32) | 0.069 |
| cg11375102 | 1.54 | (0.94, 2.50) | 0.084 |
| cg02193187 | 1.44 | (0.95, 2.19) | 0.084 |
| cg26390081 | 1.43 | (0.93, 2.19) | 0.106 |
| cg08296037 | 1.48 | (0.88, 2.51) | 0.143 |

\*Results for B-cells were not statistically significant and thus are not shown.
<sup>†</sup>Adjusted for age at blood draw, race, date of blood draw, cohort study, smoking status, and BMI.

### GSEA analysis

Using a GSEA analysis [16], we identified 225 pathways that overlapped in the two datasets (training and testing). After multiple comparisons adjustment, four pathways were borderline statistically significant in the training dataset (p-values = 0.056), and 14 pathways were statistically significant in the testing dataset (p-values<0.05) (Supplemental Table 5). The two overlapping pathways between the top findings in two datasets were gastric acid secretion and melanogenesis. The gastric acid secretion pathway includes 72 genes; the normalized enrichment score was 1.258 (testing) and 1.127 (training). The melanogenesis pathway includes 100 genes; the normalized enrichment score was 1.175 (testing) and 1.100 (training).

Using cell specific analyses, we examined pathways using GSEA in all cases and controls; none of the cell specific pathways were significant after correction for multiple comparisons, with the exception of neutrophils (Supplemental Table 6). For neutrophils, 5 pathways were statistically significant (p-value <0.05) after a Bonferroni adjustment; type I diabetes, acute myeloid leukemia and endometrial cancer pathways were among those. Of note, “pancreatic cancer” pathways were identified in NK cells (p-value = 0.007 prior to multiple comparison correction).

## Discussion

In this large study, pooling pancreatic cancer cases from 3 cohort studies, we report an association between a region of chromosome 16 (near genes *TMEM204* and *IFT140*) and risk of pancreatic cancer; the CpGs identified in this region were located primarily in a CpG island and were hypermethylated in cases compared to controls. On average, we observed higher DNA methylation levels in this region in subjects who had blood drawn closer to time of cancer diagnosis, suggesting that DNA methylation in this region, as assessed in the whole blood array platform, increases as disease progresses. Furthermore, associations between methylation and pancreatic cancer risk in CpGs located in the identified region were stronger in lymphocyte subtypes, specifically CD4 and CD8T cells, than in myeloid cells, suggesting that this could result from shifts in the percentages of smaller activated subtypes of lymphocytes. We also identified two pathways where methylation levels were related to risk of pancreatic cancer, namely gastric acid secretion and melanogenesis.

Several of the probes in the DMR region on chromosome 16 we identified have been previously identified as significant in carcinogenesis, including in pancreatic cancer. In the TCGA data, DNA methylation levels of probe cg08296037 were strongly inversely correlated with expression of TMEM204 in peripheral blood of acute myeloid leukemia patients (r = -0.79; p-value 2.3 × 10^−39^; http://gdac.broadinstitute.org/runs/analyses__2013_01_16/reports/cancer/LAML-TB/correlate/methylation_vs_mRNA/nozzle.html). TMEM204 is expressed in all cancers with low specificity, but mean levels are higher in pancreatic cancer tissue than any other cancer tissue (RNAseq TCGA data); low TMEM204 expression has been associated with poor liver cancer outcome but improved survival in melanoma (https://www.proteinatlas.org/ENSG00000131634-TMEM204/pathology).

In addition, higher levels of DNA methylation in cg09757087 were noted in a study examining the impact of insulin on DNA methylation levels in skeletal muscle of 10 healthy sedentary men with normal glucose tolerance (comparing insulin incubated muscle biopsies to basal level; paired p-value=0.006)[18]. Based on this finding, we searched GEO datasets to examine whether DNA methylation in blood varied by insulin resistance or diabetes status. In a study of 80 obese women, DNA methylation levels at cg08296037 and other probes in that region were consistently higher in peripheral blood of women with high insulin sensitivity compared to those with low insulin sensitivity (beta methylation difference range 0.04-0.06, p-value=0.07 for cg08296037; GSE76285[19]). In a study of 63 subjects with type I diabetes, methylation levels were in the opposite direction in probes in the region of interest for subjects experiencing diabetes-related progression, but the differences were not statistically significant (GSE76169)[20]. In our data, methylation levels in the DMR on chromosome 16 were lower in subjects (controls) with diabetes than those without (data not shown), which appears to conflict with the positive association between diabetes and pancreatic cancer. However, it is possible that variations in DNA methylation levels are influenced by changes in insulin levels and that treatment in individuals with diabetes alters methylation levels in that region. While it is unlikely that this region could be used to identify those at higher risk, better understanding of what influences methylation in this region and how this is impacting carcinogenic pathways may provide important clues into the etiology of pancreatic cancer.

The findings from the GSEA analysis suggest that methylation changes in the gastric secretion pathway may play a role in pancreatic cancer. Ulcers have been linked to pancreatic cancer in several studies [21-23], especially for gastric ulcers, and while these associations increase as the time to diagnosis is shorter, they are present up to 10 years prior to cancer diagnosis [22]. It has also been suggested that stomach acid secretion may alter bacterial landscape in the stomach and influence nitrosamine levels that, jointly, may contribute to pancreatic carcinogenesis [24]. Identification of changes in DNA methylation in the gastric acid secretion pathways (in cases vs controls) may provide new insights into biological mechanisms; other studies will have to confirm and further examine these findings.

Positive results from this study should be interpreted with caution given that some findings may have been due to chance; however, we tried to minimize chance findings by splitting our data into a training set and a testing set and emphasizing the results that were consistent in both sets. On the other hand, differences in methylation levels between cases and controls in single CpGs might have been missed because of the large number of corrections for multiple comparisons (>800,000). Our multivariate models adjusted for age, sex, race, smoking, and BMI to control for potential confounding, but we did not adjust for medical conditions that are known to increase the risk of pancreatic cancer, such as chronic pancreatitis, as we did not have those data available in our dataset. Finally, because we had a limited number of non-Caucasian participants in this study, our findings may not be generalizable to other populations.

To our knowledge, this is the first study to conduct an epigenome-wide association study (EWAS) on pancreatic cancer risk using peripheral blood collected prior to cancer diagnosis. Blood samples were collected an average of 13 years prior to cancer diagnosis, thereby reducing chances that the results were driven by the cancer itself. By splitting our dataset into a training and testing set, we identified a region in chromosome 16 that was differentially methylated in subjects who later developed pancreatic cancer. We also found that genes involved in gastric secretion were differentially methylated in cases compared to controls.

Findings from this study suggest that changes in methylation levels that occur more than 10 years prior to cancer diagnosis can influence risk and may be markers of altered pathway involved in carcinogenesis. For some CpGs in this region, methylation differences increased in magnitude closer to the time of diagnosis, perhaps reflecting changes in metabolic systems, a pattern that is similar to that observed between diabetes and pancreatic cancer risk. While differences in methylation levels may appear to be modest, it is important to remember the progress made and insights gained from genome-wide associations studies, even with small effects sizes [25, 26]. Results from epigenome-wide association studies (EWAS) using peripheral bloods to examine associations with cancer risk are currently sparse, but additional studies will unquestionably provide critical insights into biological pathways that could lead to major breakthroughs in clinical settings.

## Data Availability

All data from this study have been deposited in dbGAP and will be available on January 3, 2020 [“DNA Methylation Markers and Pancreatic Cancer Risk in 3 Cohort Studies (NHS, PHS, HPFS)” phs001917.v1.p1].

https://www.ncbi.nlm.nih.gov/projects/gapprev/gap/cgi-bin/study.cgi?study_id=phs001917.v1.p1

## Data Availability

https://www.ncbi.nlm.nih.gov/projects/gapprev/gap/cgi-bin/study.cgi?study_id=phs001917.v1.p1

## Data Availability Statement

All data from this study have been deposited in dbGAP and will be available on January 3, 2020 [“DNA Methylation Markers and Pancreatic Cancer Risk in 3 Cohort Studies (NHS, PHS, HPFS)” phs001917.v1.p1]. https://www.ncbi.nlm.nih.gov/projects/gapprev/gap/cgi-bin/study.cgi?study_id=phs001917.v1.p1

